# The impact of the COVID-19 pandemic on gastrointestinal infection trends in England, February – July 2020

**DOI:** 10.1101/2021.04.06.21254174

**Authors:** Nicola K. Love, Alex J. Elliot, Rachel M Chalmers, Amy Douglas, Saheer Gharbia, Jacquelyn McCormick, Helen Hughes, Roger Morbey, Isabel Oliver, Roberto Vivancos, Gillian Smith

**Author notes:** Correspondence to Nicola K Love.

## Abstract

**Objective:** To establish the impact of the first six months of the COVID-19 outbreak response of gastrointestinal (GI) infection trends in England.

**Design:** Retrospective ecological study using routinely collected national and regional surveillance data from eight Public Health England coordinated laboratory, outbreak and syndromic surveillance systems using key dates of UK governmental policy change to assign phases for comparison between 2020 and historic data.

**Results:** Decreases in GI illness activity were observed across all surveillance indicators as COVID-19 cases began to peak. Compared to the 5-year average (2015-2019), during the first six months of the COVD-19 response, there was a 52% decrease in GI outbreaks reported (1,544 vs. 3,208 (95% CI: 2,938 – 3,478) and a 34% decrease in laboratory confirmed cases (27,859 vs. 42,495 (95% CI: 40,068 – 44,922). GI indicators began to rise during the first lockdown and lockdown easing, although all remained substantially lower than historic figures. Reductions in laboratory confirmed cases were observed across all age groups and both sexes, with geographical heterogeneity observed in diagnosis trends. Health seeking behaviour changed substantially, with attendances decreasing prior to lockdown across all indicators.

**Conclusions:** There has been a marked change in trends of GI infections in the context of the COVID-19 pandemic. The drivers of this change are likely to be multifactorial; while changes in health seeking behaviour, pressure on diagnostic services and surveillance system ascertainment have undoubtably played a role there has likely been a true decrease in the incidence for some pathogens resulting from the control measures and restrictions implemented. This suggests that if some of these changes in behaviour such as improved hand hygiene were maintained, then we could potentially see sustained reductions in the burden of GI illness.

**Strengths and limitations of this study:**

- Our findings show that there has been a marked change in the burden of GI infections during the COVID-19 outbreak, and although undoubtably changes to health care and surveillance ascertainment have played a role, there does appear to be a true decrease in incidence. These findings suggest that if effective implementation of infection control measures were maintained, then we could see sustained reductions in the person to person transmission of GI illness in England.
- This study was strengthened by the triangulation of data from several national and regional-based surveillance systems; using this approach we could determine that the trends observed were consistent across all indicators.
- It has not been possible to definitively differentiate the relative contributions of the reduced ascertainment of GI infections versus a true decrease in GI disease burden in this study, which an additional focussed analysis could address.
- This analysis includes only the first six-months of the COVID-19 outbreak response, and further longitudinal analyses will be performed to explore this further and assess any change as we move into further phases of the pandemic

## Introduction

The 2019 coronavirus pandemic (COVID-19) has resulted in unparalleled challenges for society [1]. During 2020, the United Kingdom (UK) Government implemented a stepwise series of public health measures designed initially to contain, and then delay transmission of the virus (Figure 1). These measures ranged from public health information campaigns, rapid identification of cases and their contacts and isolation of those contacts in the initial phase of the outbreak (February to Mid-March 2020), with additional measures such as social distancing, education and business closures and enforceable ‘lockdown’ measures in the delay phase (Mid-March onwards; [2]). Changes to healthcare provision and patient management were implemented concurrently to alleviate pressure on the National Health Service (NHS) and minimise nosocomial transmission of COVID-19 while continuing to provide essential care [2].

**Figure 1.**
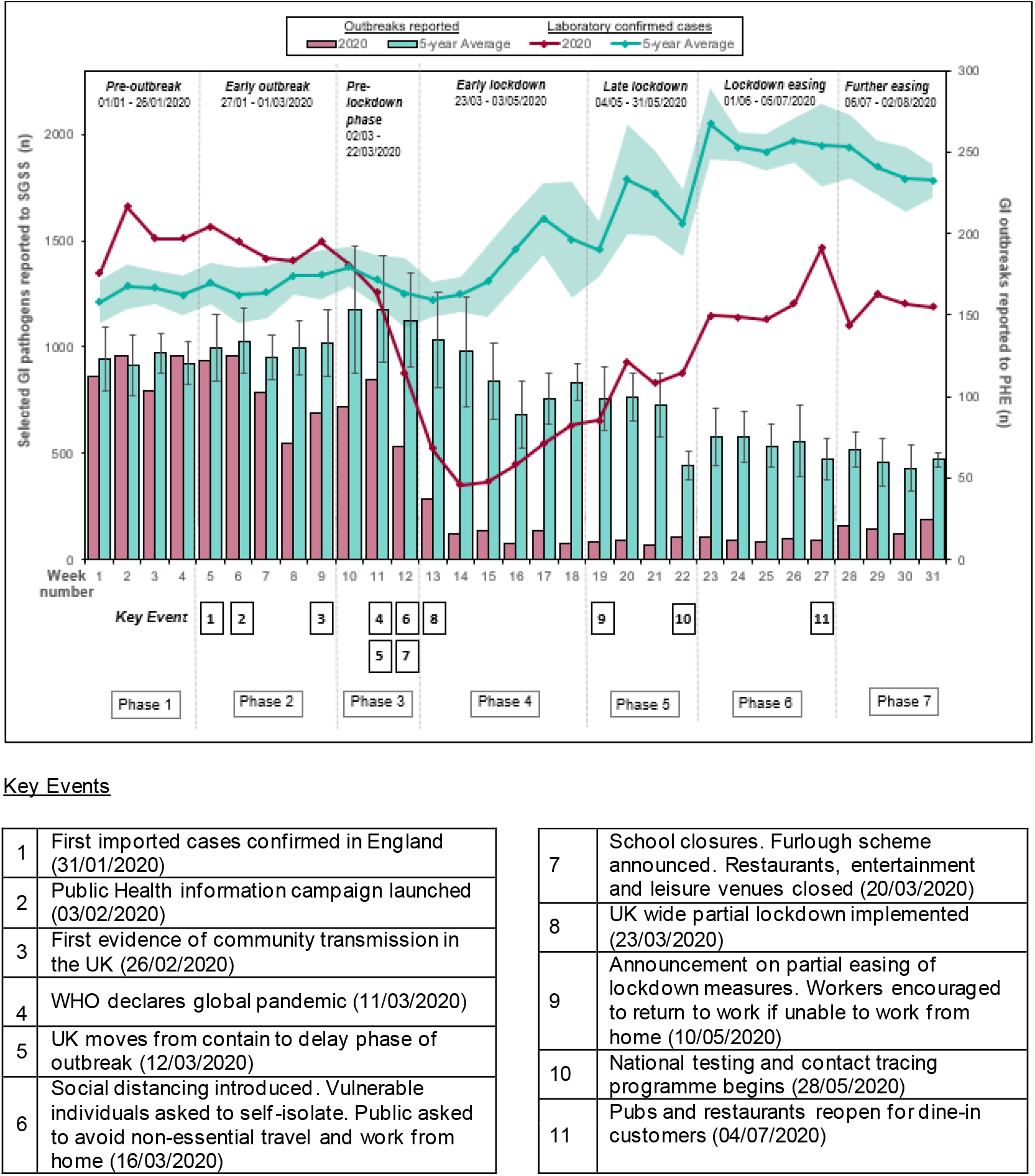
Gastrointestinal (GI) outbreaks and GI pathogens^≠^ reported to Public Health England between Week 1 and Week 31 2020 and the 5-year weekly average (95% confidence interval), indicating key public health measures introduced during the COVID-19 response

There is a growing body of evidence indicating that the COVID-19 pandemic and implemented control measures have had indirect impacts on other health conditions. Substantial decreases have been observed in emergency department (ED) attendances, with decreasing presentation of conditions such as strokes, surgical emergencies and cardiac emergencies, delays to cancer diagnoses, and concerns raised about delayed presentation and associated negative outcomes [3-7]. Less well documented are any indirect effects on communicable diseases, which are often controlled using similar non-pharmaceutical interventions to those implemented in the COVID-19 response [8].

Gastrointestinal (GI) infections are an important infectious cause of morbidity and mortality globally, placing a considerable burden on primary and secondary healthcare services. In England, it is estimated that there are in excess of 17 million cases annually, resulting in over 1 million healthcare consultations and around 90,000 confirmed laboratory diagnoses [9-10]. Transmission of GI pathogens is typically faecal-oral, predominantly though consumption of contaminated food or water, or contact with infected individuals, animals or the contaminated environment and fomites; importance of transmission route varies substantially by pathogen. Control measures implemented during the COVID-19 response including improved hand hygiene, reduced social contact, increased environmental cleaning and closure of premises, all known to be effective in reducing GI infections, primarily those spread by person-to-person transmission and environmental contamination [8].

Using routinely collected surveillance data from several English surveillance systems coordinated by Public Health England (PHE), this study aimed to establish what impact the first six months of the COVID-19 outbreak response (27 January – 2 August) have had on trends in GI infections.

## Methods

### Surveillance systems

A retrospective ecological study was conducted by performing secondary analyses on routinely collected national and regional surveillance data from eight national PHE coordinated surveillance systems, detailed in Table 1, and Google Trend data. Systems included outbreak monitoring (HPzone), laboratory notifications Second Generation Surveillance System, (SGSS) and EpiNorth3, and real-time syndromic surveillance [11-13]. Syndromic systems covered the spectrum of severity ranging from the NHS 111 telephone health advice service and routine medical appointments captured in the general practitioner (GP) ‘in hours’ system (GPIH), to GP ‘out of hours’, which covers emergency GP appointments for acute or severe illness and the Emergency Department Syndromic Surveillance System (EDSSS), which captures those attending EDs [13].

**Table 1.** PHE Surveillance systems and indicators for gastrointestinal infection surveillance

| **System** | **Coverage** | **Reporting statistic** | **Indicator** |
| --- | --- | --- | --- |
| **HPzone** | National (by PHE regional centre) | Statutory notifications of suspected or confirmed outbreaks of food poisoning, haemolytic uraemic syndrome (HUS) and infectious bloody diarrhoea.<br>Outbreaks of gastroenteritis in closed settings. | Food poisoning;<br>HUS;<br>Infectious bloody diarrhoea.<br>GI illness in closed settings (Care homes, schools, prisons) |
| **EpiNorth3** | North East England residents (approx. 2.6 million population) | Notifiable infections reported in North East England linked with enhanced exposure information from routine surveillance questionnaires conducted by environmental health officers for selected organisms. | Specified GI pathogens:<br>Cryptosporidium, Shiga toxin producing E. coli (STEC), Giardia, Salmonella, Shigella |
| **SGSS (Second Generation Surveillance System)** | All NHS laboratories in England | Routine laboratory notifications from NHS laboratories including statutory notifications of laboratory confirmed notifiable organisms. | Specified GI pathogens:<br>Campylobacter, Cryptosporidium, E. coli (STEC), Giardia, Listeria, Norovirus, non-typhoidal Salmonella, Shigella |
| **NHS 111** | National, England | Calls for indicator | Diarrhoea, Vomiting |
| **GP in hours** | Approximately 4,000 GP practices in England | In-hours (weekdays, daytime) GP consultation rates per 100,000 registered patients | Gastroenteritis – clinical diagnoses,<br>Diarrhoea and Vomiting |
| **GP out of hours** | Approximately 60 % coverage of GP out-of-hours activity in England | Out-of-hours and unscheduled care consultations for indicator as a percentage of total read-coded consultations | Gastroenteritis – clinical diagnoses,<br>Diarrhoea and Vomiting |
| **Emergency Department (ED)** | Sentinel system. 80 EDs across England | Percentage of emergency department visits with a recorded diagnosis code. | Gastroenteritis – clinical diagnoses, |

### Time periods

Extracted data covering weeks 1 to 31 2020 (30 December 2019 – 2 August 2020), and historic comparator data from weeks 1-31 2015-2019, was split into seven COVID-19 pandemic ‘Phases’ for comparison, determined by the key dates of UK governmental policy changes implemented during the COVID-19 response (Figure 1). These Phases comprised: Pre-outbreak (Phase 1); Early outbreak (Phase 2); Pre-lockdown (Phase 3); Early lockdown (Phase 4); Late lockdown (Phase 5); Lockdown easing (Phase 6); and Further easing (Phase 7; Table 2).

**Table 2.**
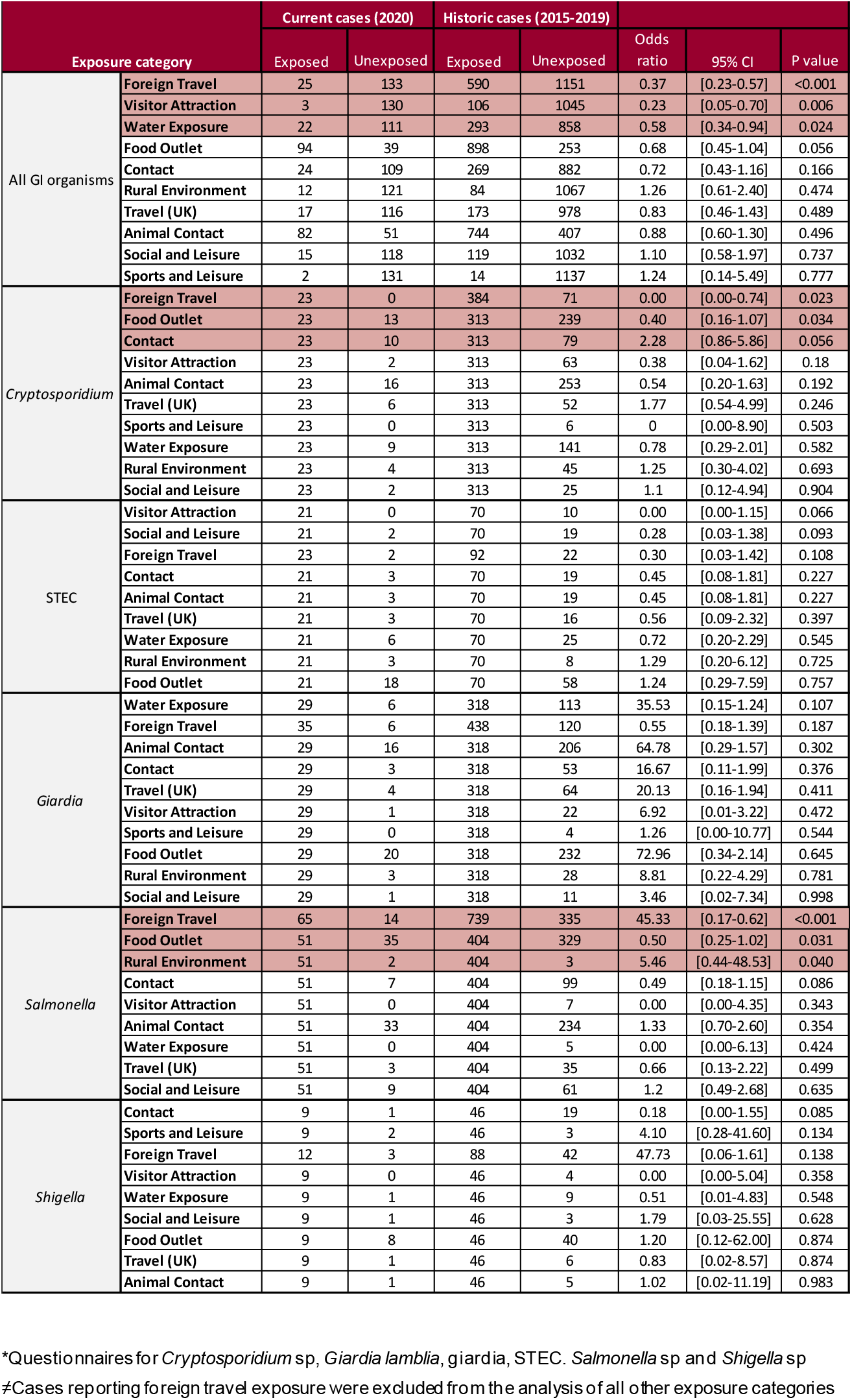
Exposures reported in routine environmental health surveillance questionnaires* for GI pathogens, North East residents. Phase 2 – Phase 7 of the COVID-19 outbreak

### Data analysis

Total weekly gastrointestinal outbreaks recorded in HPZone were determined for the seven Phases of 2020 and the 5-year average (2015-2019) with 95% confidence intervals calculated. HPZone data were further analysed by outbreak setting, PHE region and pathogen (including if the suspected pathogen was laboratory confirmed). Pseudonymised SGSS data for selected laboratory confirmed organisms (*Campylobacter* spp., *Cryptosporidium* spp., Shiga-toxin producing *E. coli* [STEC], *Giardia* sp., *Listeria* spp., Norovirus, non-typhoidal *Salmonella* spp., *Shigella* spp.) were grouped by week of specimen sample date. Age group specific rates were calculated per 100,000 population using Office National Statistics 2019 England mid-year population estimates [14]. Cumulative regional and local authority rates per 100,000 population were determined for week 1 to 31 2020 and the 5-year average for GI infections reported to SGSS for nine PHE regions (ranging from 2.6 – 9.1 million population) and 150 local authority areas (average population size: 375,686). For geographies, risk ratios and percentage relative effects were calculated, and Pearson’s correlation was performed for each local authority area using the cumulative COVID-19 rate per 100,000 population [15].

Syndromic data were analysed as described elsewhere [16] for data between week 1 and week 31 2020, with the same period in 2019 used as a comparator. Google Trend searches were performed for key phrases associated with GI illness in England, as described previously [17]. A score out of 100 is used to represent relative search interest over the given time period and geography.

EpiNorth3 data were used to compare cases from Phases 2-7, 2020 to historic cases (2015-2019). A univariate case-case analysis was performed using exposure data, with cases reporting foreign travel excluded from analyses of other exposure categories. Comparisons were made between symptom presentation for historic and current cases. Time periods between onset date and specimen date and specimen date and referral date, and the duration of illness were determined, and a Mann-Whitney U test performed. Data from 2019 and 2020 were used to look at differences in laboratory diagnostic testing methods tests used, to account for the increasing use of molecular based techniques in recent years.

All variables were plotted in a time series together with the weekly 5-year average and superimposed COVID-19 outbreak Phases, unless otherwise specified. Statistical analyses were performed using Stata software version 14.2.

### Patient and Public Involvement

This study used routinely collected surveillance data. Patients were not involved in the development of the research question and outcome measures, the design of the study, the recruitment and conduct of the study.

## Results

### The changing trends of gastrointestinal infections during the different ‘Phases’ of the COVID-19 outbreak

During the first seven months of 2020, 1,544 suspected and laboratory-confirmed GI outbreaks were reported in England, representing a 52% decrease on the 5-year average for the period (3,208 (95% CI: 2,938 – 3,478)). During the ‘Pre-outbreak’ (Phase 1; weeks 1-4), notified GI outbreaks were comparable to historic figures (Figure 1 and Supplementary Table 1). From week 7 (‘Early outbreak’, Phase 2) there was a 22% decrease in GI outbreaks (510 outbreaks vs. 5-year average: 651 (95% CI: 605-697), with this decreasing trend continuing to an 87% reduction in GI outbreaks during ‘Late lockdown’ (Phase 5; weeks 19-22; 5-year average: 46 vs. 350 outbreaks (95% CI: 294 – 406). Reported outbreaks remained substantially lower than historically observed for the duration of the COVID-19 response period.

Historically, around 95% of suspected or confirmed outbreaks reported in England are attributed to viral GI pathogens (94% in 2020; Supplementary Table 1) primarily occurring in health and social care settings. During the COVID-19 response period (Phases 2-7), there was a 62% reduction in the number of reported suspected and confirmed viral outbreaks (862 vs. 2,239), with significant reductions in parasitic outbreaks (2 vs. 32 (95% CI: 21-44); 94% decrease) and bacterial outbreaks (47% decrease; 51 vs. 97 outbreaks (95% CI: 77-115)). Significant decreases in laboratory confirmed GI cases also occurred, with 27,859 cases reported between Phases 2 and 7 compared with a 5-year average of 42,495 (95% CI: 40,068–44,922; 34% decrease; Figure 1 and Supplementary Table 1). Decreased reports were apparent from week 10 (‘Pre-lockdown’; Phase 3; 11% decrease), with a low of 2,859 cases reported between weeks 13-18 (‘Early lockdown’; Phase 4) representing a 66% decrease (5-year average: 8,345; (95% CI: 7,602-9,088)). Laboratory confirmed cases began to increase from week 16 onwards, mirroring the historic seasonal trend for reported GI pathogen activity despite numbers remaining significantly lower than average; during the historic peak for laboratory reporting, which occurred during lockdown easing, 4,617 cases were reported compared with the 5-year average of 7,879 (95% CI: 7,539-8,219); 41% decrease).

While the total number of laboratory-confirmed cases was reduced, causative organisms were differently impacted when compared to the five-year average. Norovirus reports were most reduced (5.6% of all laboratory confirmed reports vs. 9.0%), with reductions also observed for *Salmonella* spp. (7.9% vs. 9.5%) and *Cryptosporidium* spp. (2.8% vs. 3.9%). The proportion of laboratory-confirmed cases with *Giardia* spp. (5.4% vs. 5.2%), STEC (1.2% vs. 1.2%) and *Listeria* spp. (0.2% vs. 0.2%) during the outbreak period remained comparable, while the proportion of *Campylobacter* spp. reports were increased (74% vs. 68%).

Reported norovirus cases decreased substantially during Phases 3 and 4 (weeks 11-14; Figure 2a), remaining significantly below the 5-year average to week 31. A similar trend was observed for *Cryptosporidium* spp., which contrasted with historic trends where a spike in activity was observed between weeks 14 and 18 (Phase 4; 89 vs. 477 cases (95%CI: 323-631); Figure 2b). The bacterial pathogens *Shigella* spp., *Salmonella* spp. and *Campylobacter* spp., all showed significant decreases during Phase 3, remained low during Phase 4 but then began to increase during Phase 5, following the 5-year average but with a significantly reduced number of cases reported (Figure 2d,f,g). The substantial decreases observed for other pathogens were not seen for STEC (Figure 2e), with activity remaining below or at the lower limit of the 5-year average.

**Figure 2.**
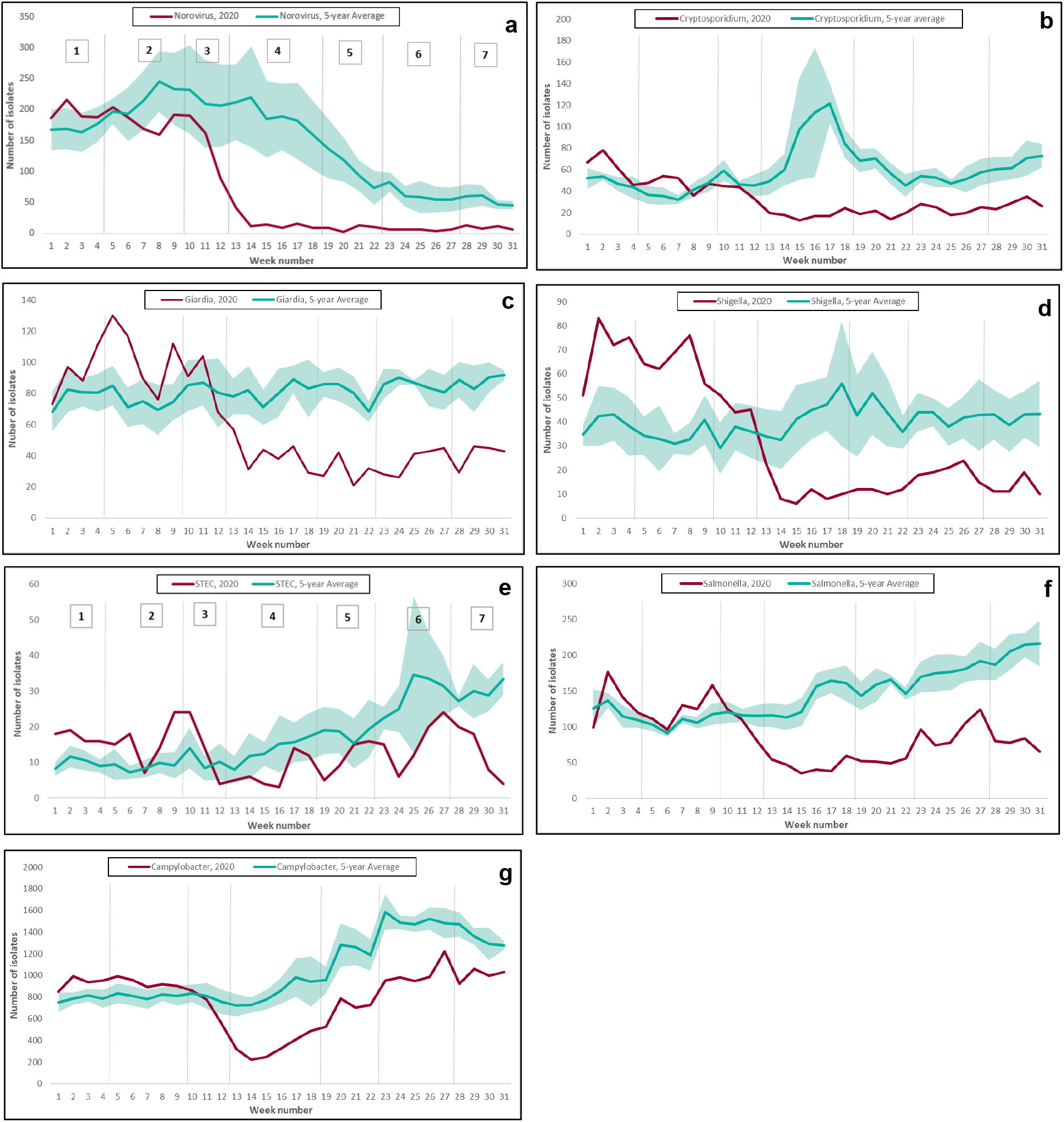
Laboratory confirmed GI pathogens reported to Public Health England between Week 1 and Week 31 2020 and the 5-year weekly average (95% confidence interval), by COVID-19 outbreak Phase

Reductions in laboratory confirmed cases across all pathogens were observed across all age groups and both sexes, with decreases varying from 26% in children aged 1-9 years to 42% in females over 80 years (Supplementary Figure 1). Age-specific rates for all age groups sharply decreased during the ‘Pre-lockdown’ (Phase 3; weeks 10 - 12) with lows observed during ‘early lockdown’ (Phase 4; weeks 13 – 18; Figure 3). Reporting rates began to increase for all age groups into the Late lockdown period (Phase 5; weeks 19-22), with infection rates in elderly adults and children under 14 years comparable to the 5-year average by week 31.

**Figure 3.**
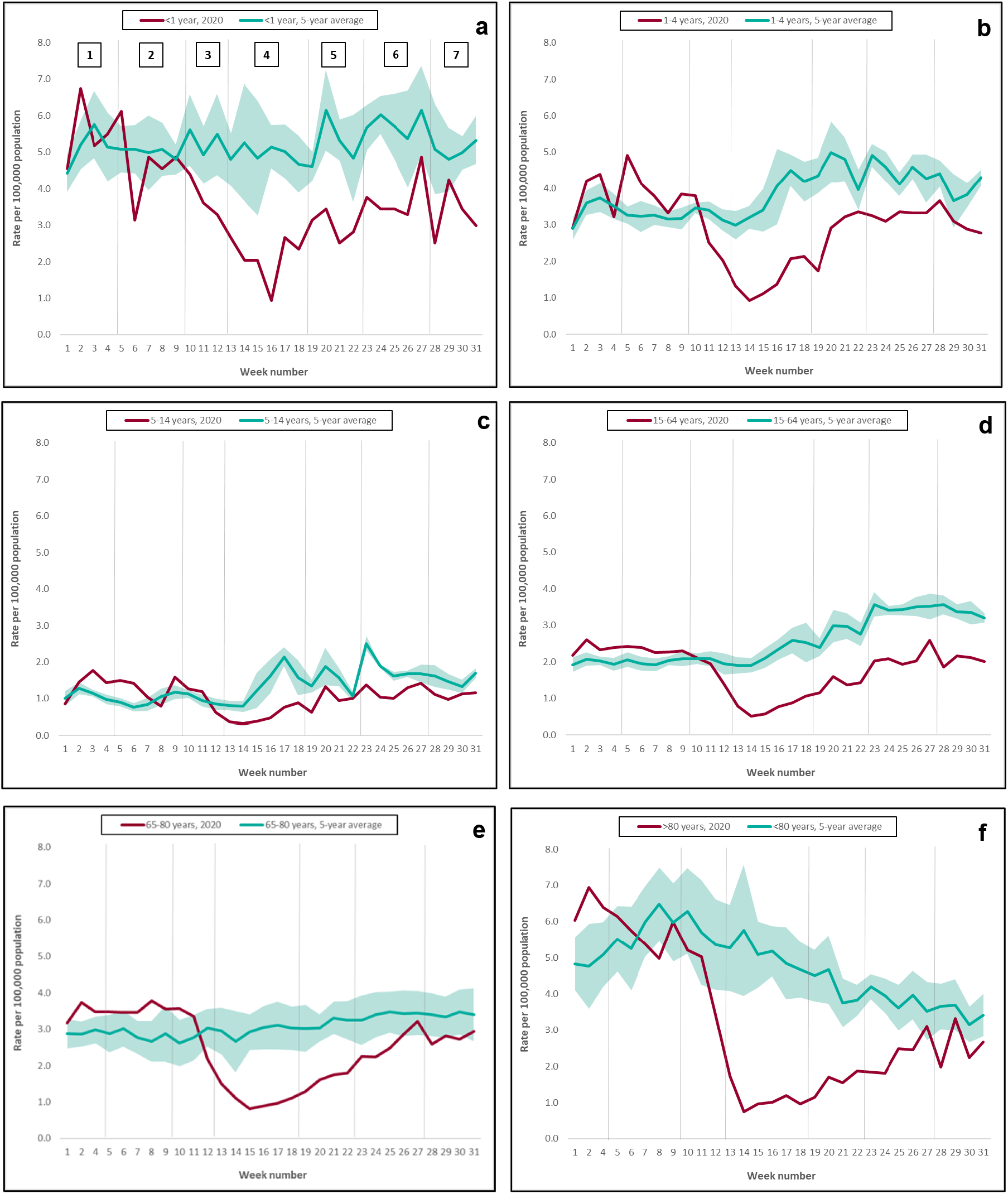
Laboratory confirmed GI pathogen^≠^ cases reported to Public Health England between Week 1 and Week 31 2020 and the 5-year weekly average (95% confidence interval), by COVID-19 outbreak Phase and age group

Geographical differences in laboratory confirmed diagnoses were observed across England’s nine regional areas (Figure 4 and Supplementary Figure 2). Prior to the COVID-19 outbreak, GI laboratory-confirmed cases in all regions were above the regional 5-year average with the highest difference in the North East, London and South East regions. By the ‘pre-lockdown Phase’ (Phase 3), laboratory diagnoses had decreased below the 5-year average in all but the North East, and by lockdown (Phases 4 and 5), all regions were below historical figures. Lockdown easing (Phases 6 and 7) resulted in small increases in GI diagnoses across all regions, with the North West (NW) showing the smallest decrease in GI pathogens over the lockdown and easing periods, compared to historical figures. There was a significant correlation between upper tier local authorities showing smaller decreases in GI laboratory reports during the COVID-19 outbreak period (Phases 2-7) and those with highest COVID-19 rates up to week 31 (Pearson’s correlation 0.18, p=0.03; Supplementary Figure 3).

**Figure 4.**
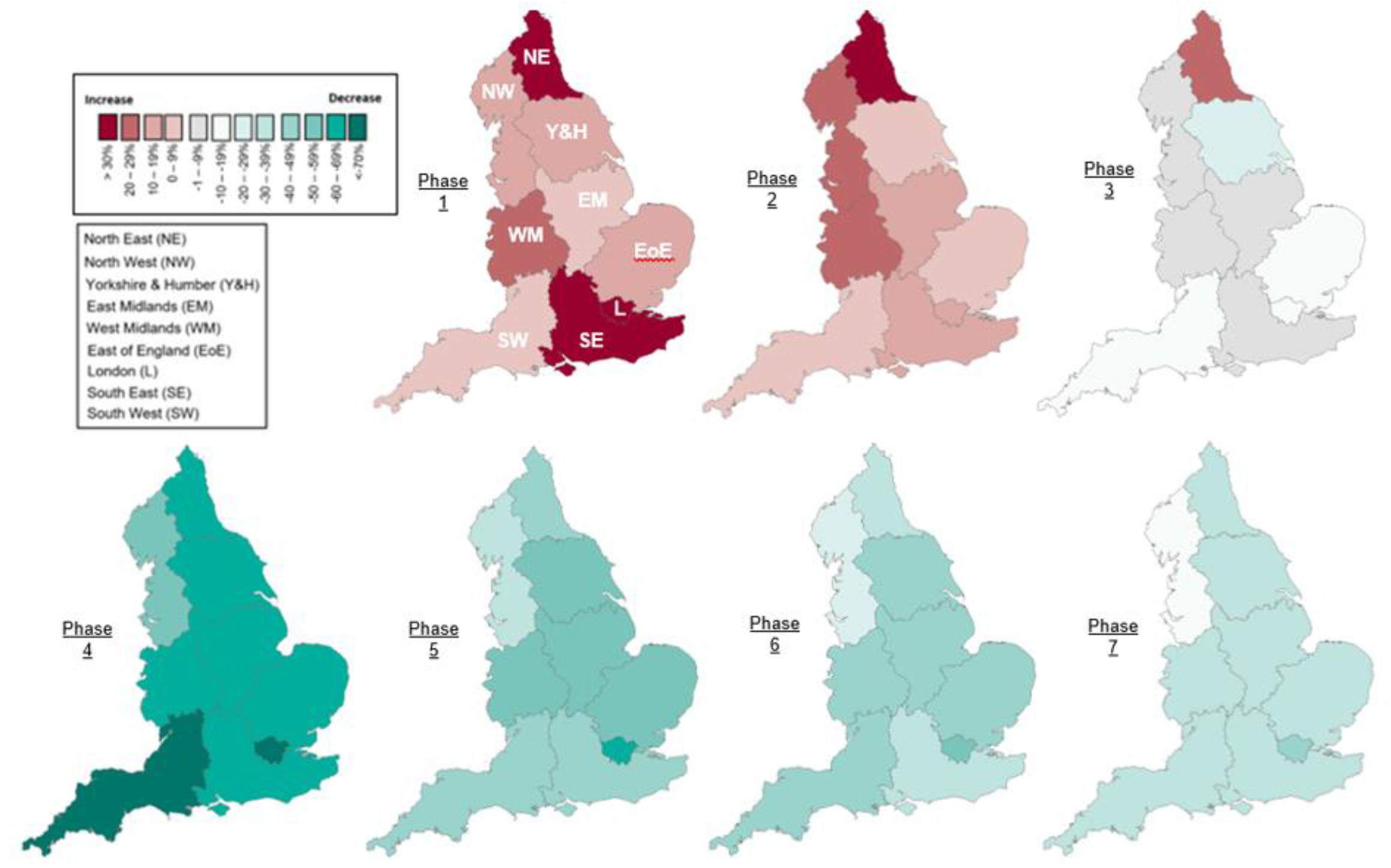
Laboratory confirmed GI cases by PHE region and COVID-19 outbreak Phase, percentage change compared to the 5-year average for the region

### The contribution of health-seeking behaviour and healthcare provision to changes in reported GI illness during the COVID-19 outbreak

Observed changes may reflect a real decrease in incidence or may be due to changes to healthcare provision or altered health-seeking behaviour or laboratory testing practices, resulting in reduced surveillance system ascertainment. ED attendances for gastroenteritis (as a proportion of attendances with a diagnostic code) decreased substantially in week 11 to a low in week 14, remaining substantially lower than the 2019 comparator up to week 31 (Figure 5a). Gastroenteritis consultations in GP-out of hours services also decreased during the ‘pre-lockdown Phase’ (Phase 3; weeks 10 - 12), increasing slightly at around week 26 but again remaining consistently low across the period (Figure 5b). A similar trend was also observed for gastroenteritis consultations to GP-in hours services (Figure 5c). Finally, calls to the NHS 111 helpline for diarrhoea and vomiting combined began to decrease from week 8 to a low at around week 12 (Figure 5d). Calls showed a gradual increase over time, remaining substantially lower than calls observed in 2019. There were no differences in the observed trends for diarrhoea or vomiting as separate symptoms (Supplementary Figure 7).

**Figure 5.**
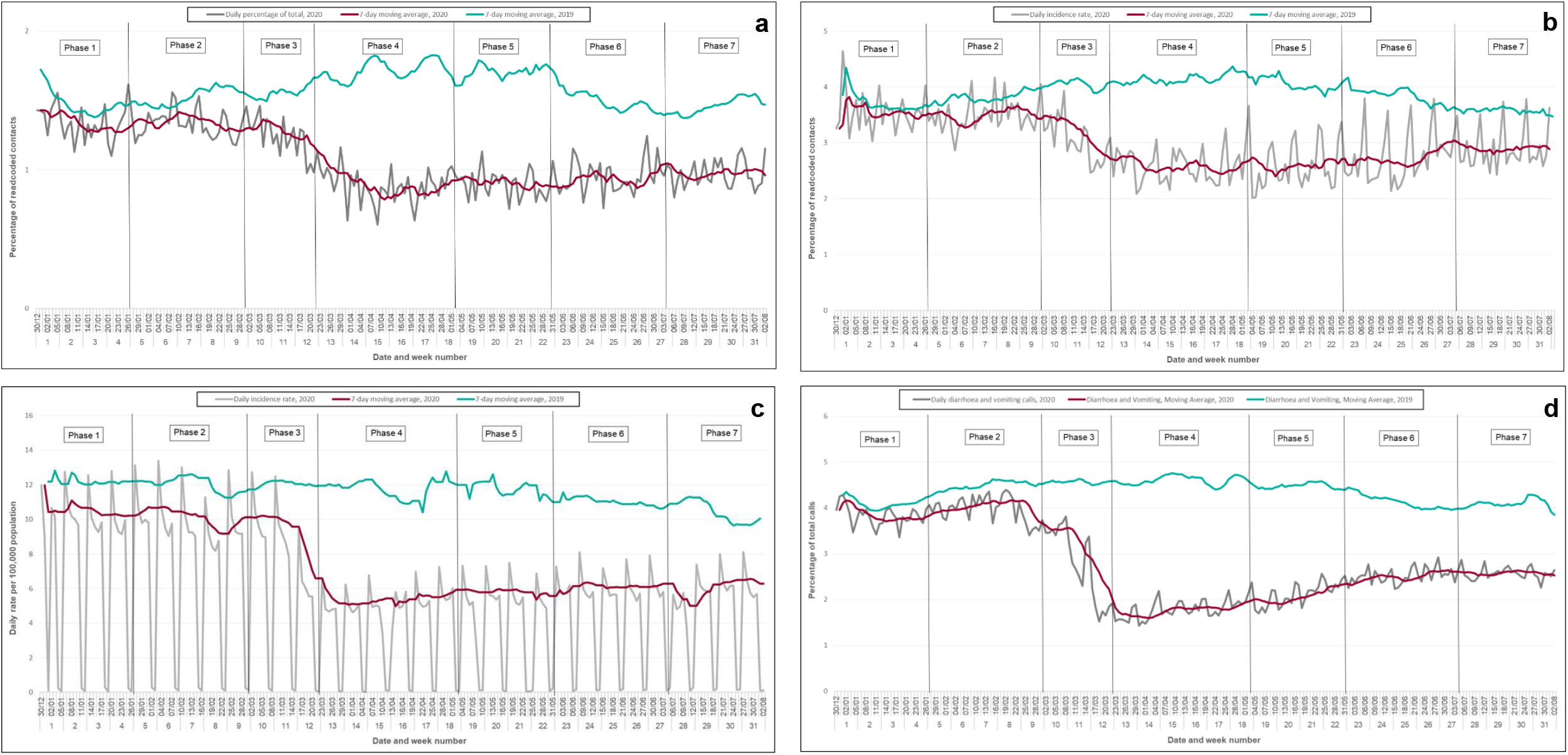
Syndromic indicators for Phases 1-7 of the COVID-19 response, 2020 and 2019

**Figure 6.**
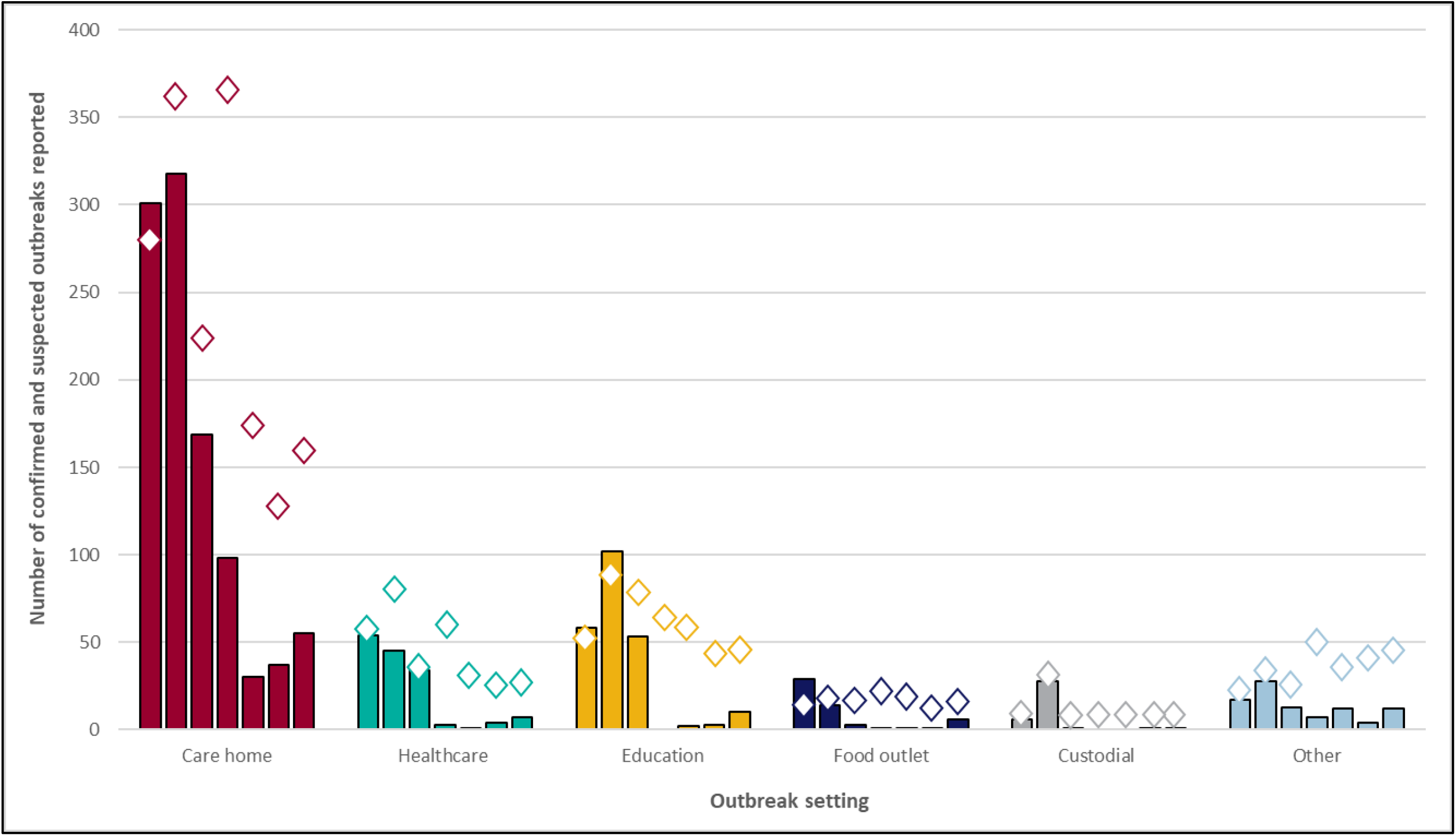
Number of gastrointestinal outbreaks reported to Public Health England by outbreak setting between Phases 1 and 7 of the COVID-19 response, with the lower bound* of the 5-year average 95% confidence interval indicated by diamonds

Decreases in GP-in hours consultations were predominantly observed in younger age groups with substantially decreased age-specific rates seen for children under 14 years during the ‘pre-lockdown Phase’ (Phase 3), and rates remaining low to week 31 (Supplementary Figure 5). More modest decreases were observed at week 11 for adults and the elderly, with consultation rates remaining stable during lockdown and lockdown easing. Trends for PHE regions (rate per 100,000 population) were comparable, with decreases for all regions in the ‘pre-lockdown Phase’ and slight increases during lockdown easing (Supplementary Figure 6).

Enhanced specimen and exposure information was obtained from EpiNorth3 for laboratory confirmed North East cases. During the outbreak period, 228 cases were reported compared with 2,225 between 2015 and 2019 (74% and 80% with a recorded onset date, respectively). There was no difference in the time between onset date and specimen date for cases reported in Phase 2-Phase 7 2020, compared with historic cases (7 days IQR: 3-14 vs. 7 days (4-12 days); Wilcoxon rank sum p=0.600), with onset to specimen time comparable across the seven Phases of the COVID-19 outbreak. The time between specimen date and referral date and proportion of samples processed by culture (76.5% vs. 75.7%) and molecular techniques (17.2% vs. 15.5%) were also comparable. However, the proportion of specimens processed by light microscopy (protozoa) decreased (2.3% vs. 7.6%). The proportion of specimens submitted by general practitioners was 60.2% compared with 61.3% historically, while a small decrease was observed in specimens from EDs (5.1% vs. 8.4%) and a small increase from hospital inpatients (29.6% vs. 25.2%).

Analysing EpiNorth3 data it was possible to explore illness duration and symptoms for cases with a completed surveillance questionnaire (n=179 [2020]; n=1870 [2015-2020]). The proportion of current cases and historic cases reporting diarrhoea (40% vs.38%) and vomiting (39% vs. 42%) were comparable, with an increase in cases reporting bloody diarrhoea in 2020 (24% vs. 16%). When comparing 2020 to historic cases, there was no difference in the duration of illness (7.5 days IQR: 5-12 vs. 8 days (6-12 days); Wilcoxon rank sum p=0.980), however as most cases are symptomatic when interviews are performed (∼26% of current and historic cases), completion of this field is low.

### The contribution of population behavioural change to impacts of COVID-19 on GI illness

Using EpiNorth3 data it was possible to look at changes in exposures for cases of GI illness reported in North East England during the COVID-19 outbreak period and historically (Phase 2-7; 2020: 79% cases reported exposure; n=230 vs. historic cases: reported between 2015 and 2019; 84% cases reported exposure; n=2,501). The odds of cases with a GI diagnosis reporting foreign travel were significantly reduced in 2020 (OR: 0.37; (95% CI: 0.23-0.57); p=<0.001), and there were also significant decreases in the odds of cases reporting exposures to UK visitor attractions (OR: 0.23; (95% CI: 0.05-0.70); p=0.006) and recreational water exposure in the UK, which includes swimming pools (OR: 0.58; (95%CI: 0.34-0.94); p=0.024). *Salmonella* and *Cryptosporidium* cases had significantly lower odds of reporting UK food outlet exposures (OR: 0.5 and 0.4 respectively, p=0.03 for both).

Trends in outbreak reporting could also be attributed in part to behavioural change. Significant reductions in health and social care outbreaks caused by all pathogens were observed from the ‘early pandemic Phase’ (Phase 2) of the COVID-19 response, with 318 care home outbreaks (vs. 5-year average: 402 (95% CI: 355-448)) and 45 healthcare setting outbreaks reported (vs. 89 (95% CI:75-103); Figure 5). In educational settings, the number of outbreaks was comparable to the 5-year average in Phase 2, with a decrease observed in Phase 3, prior to school closures (53 outbreaks vs. 86 (95% CI: 70-102)). Outbreaks in educational settings remained low following the reopening of schools for specific age groups (∼25% of pupils) in week 23 (Phase 6; 3 vs. 61 outbreaks (95% CI: 37-84)), with the number of outbreaks increasing during Phase 7 (10 vs. 51 outbreaks (95% CI: 39-64)). Outbreaks associated with food outlets also reduced significantly in Phase 3, prior to the lockdown period (3 vs. 13 (95% CI: 9-17)) and remained low until Phase 7 when pubs and restaurants reopened for dine-in customers (6 vs. 13 (95% CI: 9-17)). Outbreaks classified as ‘other’, which included visitor attractions, were substantially decreased in Phase 4, when outbreaks associated with such settings are historically highest (7 vs. 51 (95% CI: 44-57)).

Finally, evidence from Google Trends data showed searches for GI associated phrases such as ‘Food poisoning’, ‘gastroenteritis’ and ‘sickness bug’ all decreased dramatically between week 11 and week 13, while trends for ‘handwashing’ and ‘disinfection’ increased substantially between week 8 and week 14, mirroring decreases observed in other surveillance systems (Supplementary Figure 8).

## Discussion

Analysis of routinely collected surveillance data from England’s main laboratory, outbreak and syndromic surveillance systems showed marked changes in GI infection trends during the first six months of the COVID-19 outbreak response. Decreases in GI activity were observed across all surveillance indicators as restrictions were implemented in line with increasing COVID-19 activity [15]. As COVID-19 cases increased and further restrictions were implemented, health-seeking behaviour changed dramatically in England, with health service attendances decreasing from week 12 across all syndromic indictors [1, 18-22]. Similar decreases were observed internationally, with speculation that reduced healthcare usage was due to public avoidance, with some evidence that those with milder symptoms were least likely to seek care [3-7, 23]. However, behavioural surveys conducted during late lockdown and lockdown easing found 70-80% of participants had continued to seek care, if needed [24-25]. Protecting the health system was cited as a reason for healthcare avoidance by 16% of respondents with symptoms, while 10% of individuals reported difficulties in accessing their GP for physical complaints [24-25]. In England, a government information campaign was launched from week 17 to encourage the public to continue to seek care, particularly for severe or acute conditions [2]; however, our study shows that GI activity was increasing prior to this. Many syndromic indicators, such as those for non-infectious gastrointestinal conditions, rapidly returned to baseline levels during lockdown and easing, while gastrointestinal infections showed more modest increases [18-21].

Behavioural studies undertaken during the first wave suggest good adherence to hygiene and social distancing measures by the population [25, 26], raising the possibility that widespread infection prevention and hygiene measures implemented during the COVID-19 response have had protective effects to varying degrees dependant on the pathogen and transmission route of GI pathogens. Indeed, evidence suggests that non-pharmaceutical interventions implemented in the response were associated with reductions in influenza activity during the 2019/20 season [27-29]. As with influenza, viral gastroenteritis activity is higher during winter, with outbreaks predominantly associated with person-to-person transmission in health and social care settings; in the 2019/20 season, this outbreak activity appeared to have been partially curtailed by the COVID-19 response. Enhanced infection control measures were recommended in closed settings during week 8 [2]; although there was evidence that measures were not being sufficiently implemented until later in the response [30]. Reports of outbreaks in food outlet and education settings were also reduced prior to closures announced as part of social distancing measures, and all surveillance indicators remained significantly lower than average despite the partial reopening of schools, restaurants and overseas travel corridors during the lockdown easing phase [2].

There was evidence that those pathogens spread predominantly by person-to-person transmission, such as norovirus and *Shigella* spp., showed greater reductions, while bacterial pathogens, which are more commonly foodborne and therefore less influenced by hygiene and social distancing measures, were least impacted [8, 31]. It is possible that those individuals with the most severe and prolonged infections, which tend to be bacterial, were more likely to access care, as seen with other diseases [4,6,8,9]. However, a true decrease in activity was plausible given the measures implemented. *Salmonella* activity was likely substantially reduced by government guidance on non-essential foreign travel, in place from week 12 [2, 32]. Campylobacter, which is usually foodborne and often associated with incorrect food preparation was less impacted than other GI pathogens, although possible explanations for reductions may include food business closures and improved hygiene limiting the risk of cross-contamination. Cryptosporidiosis has strong seasonality with two peaks, one in late spring associated with *C. parvum* which occurs around lambing season and is often associated with petting farm outbreaks, which were closed as part of the COVID response, and the other, usually larger peak in late summer-early autumn mainly caused by *C. hominis* linked to increased recreational water use and foreign travel [33]. However, in 2020, *C. hominis* cases were virtually absent (Cryptosporidium Reference Unit data). It is likely that the closure of premises such as swimming pools and open farms have played a considerable role in this decline, and their reopening may impact on disease transmission.

Reasons for the changes in the national picture observed for GI infections are likely to be complex and multifactorial, and it is not possible to attribute them to a specific cause. This study was strengthened by the triangulation of data from several surveillance systems; using this approach we could determine that the trends observed were consistent across all indicators. However, while this study has incorporated large national and regional-based surveillance systems, this is not comprehensive and there are other examples of operational GI surveillance systems which have not been included in this study. For example, the PHE eFOSS system monitors foodborne and non-foodborne GI outbreaks across England, however, due to a small number of outbreaks reported by the system it was felt that these data were not sufficiently powered to add to the overall findings of the study [34]. There are further limitations to this work; it has not been possible to definitively differentiate the relative contributions of the reduced ascertainment of GI infections versus a true decrease in GI disease burden in this study, which an additional focussed analysis could address. An additional limitation of this study is that negative results are not captured by the SGSS laboratory surveillance system, therefore it was not possible to determine to what degree the changes were due to changes in testing. We were not able to calculate the positivity rate to assess whether only severe cases were being tested, although evidence from the North East suggested that symptomatology was comparable between 2020 cases and historic cases.

Guidance was released in week 13 recommending the cessation of routine culture for non-bloody diarrhoea might be considered if laboratories were struggling to deliver the service [35]. However, by then laboratory confirmed cases were beginning to increase again, while other contemporaneous changes implemented to GI laboratory testing methods, such as the introduction of molecular methods could differentially affect ascertainment of GI pathogens. Furthermore, the smallest decreases in laboratory confirmed GI infections were observed in the North West and North East regions, which had the highest cumulative rates of COVID-19 over this time period [15], suggesting either increased laboratory capacity for both the identification of COVID-19 and GI infections, or less effective implementation of COVID-19 control measures resulting in greater spread of GI infections and COVID-19.

This comprehensive review of PHE surveillance data demonstrates a marked change in GI infection trends in the context of the COVID-19 epidemic. The change is likely to be multifactorial; changes to health seeking behaviour have undoubtedly played a significant role in the trends observed, and ascertainment through surveillance systems has likely been affected. However, from our observations it appears that there has probably also been a true decrease in the incidence of GI infections most likely associated with control measures for the COVID-19 epidemic, although the extent of the true decrease cannot be fully estimated in light of the multiple influencing factors identified and the variation in the ecology and epidemiology of GI pathogens and transmission routes. The findings of this study show the importance of multiple surveillance systems to allow for the comparative analysis across multiple indicators in order to overcome the negative impact of a pandemic situation on national surveillance systems and allow us to see the effects. Secondly, because they suggest that if measures such as improved hand hygiene and the effective implementation of infection prevention and control measures in health and social care settings were maintained, then it is possible to see sustained reductions in the burden of GI illness [30].

This analysis includes only the first six-months of the COVID-19 outbreak response, and further longitudinal analyses will be performed to explore this further and assess any change as we move into further phases of the pandemic including the relaxation of social distancing measures, further lockdown measures, and the usual winter outbreak period.

## Data Availability

No additional data available

## Contributorship statement

N.L., S.G., I.O and G.S. designed the study. N.L. analysed the data, with support from A.E., A.D., G.S., H.H., and R.M. R.C., A.D., S.G., J.M., and R.V. contributed to the interpretation of the results. N.L., A.E, and G.S. wrote the manuscript with input from all authors.

## Acknowledgements

The authors acknowledge the contribution and support from all data providers including: NHS 111 and NHS Digital; QSurveillance®, University of Oxford, EMIS/EMIS practices, ClinRisk®, TPP, ResearchOne and participating SystmOne GP practices; Advanced and the participating OOH service providers; participating EDSSS emergency departments, Royal College of Emergency Medicine; North East, North West, Yorkshire, East Midlands, West Midlands, East of England, London, South East Coast, South Central, and South Western NHS Ambulance Trusts and The Association of Ambulance Chief Executives. The authors also wish to thank Petra Manley, Katri Jalava and Lesley Larkin for comments and Claire Ferraro, Lucy Findlater and Adrian Wensley for technical support.

## Ethical approvals

Ethical approval was not required for this study. The authors affirm that the manuscript is an honest, accurate, and transparent account of the study being reported; that no important aspects of the study have been omitted; and that any discrepancies from the study as originally planned have been explained.

## Data sharing

No additional data available

