## Supplemental Figures for "The impact of the COVID-19 pandemic on gastrointestinal infection trends in England, February – July 2020"

**Supplementary Figures**

**Supplementary Table 1 – Gastrointestinal outbreaks and GI pathogens≠ reported to Public Health England between Week 1 and Week 31 2020 and the 5-year weekly average (95% confidence interval), by COVID-19 outbreak Phase**

| | Phase | 1 | 2 | 3 | 4 | 5 | 6 | 7 |
|---|---|---|---|---|---|---|---|---|
| | Week | 1-4 | 5-9 | 10-12 | 13-18 | 19-22 | 23-27 | 28-31 |
| | | Pre-outbreak phase | Early outbreak | Pre-lockdown | Early Lockdown | Late Lockdown | Lockdown Easing | Further Easing |
| Gastrointestinal outbreaks | 2020 | 465 | 510 | 273 | 109 | 46 | 50 | 91 |
| | 2019 | 545 | 684 | 383 | 752 | 443 | 326 | 314 |
| | 5 year average (95% CI) | 489 (444 - 534) | 651 (605 - 697) | 454 (357 - 551) | 667 (554 - 780) | 350 (294 - 406) | 292 (226 - 358) | 305 (273 - 337) |
| | % Change | -5% | -22% | -40% | -84% | -87% | -83% | -70% |
| Viral outbreaks | 2020 | 390 | 441 | 224 | 80 | 27 | 36 | 54 |
| | 5-year average (95% CI) | 417 (377-457) | 558 (512-604) | 391 (305-477) | 555 (449-660) | 281 (239-324) | 225 (174-276) | 229 (203-256) |
| Bacterial outbreaks | 2020 | 9 | 9 | 4 | 8 | 9 | 7 | 14 |
| | 5-year average (95% CI) | 11 (7-14) | 12 (8-15) | 7 (4-10) | 18 (16-19) | 18 (14 -22) | 18 (15-22) | 24 (20-27) |
| Parasitic outbreaks | 2020 | 2 | 1 | 0 | 1 | 0 | 0 | 0 |
| | 5-year average (95% CI) | 4 (2-5) | 3 (1-6) | 4 (2-5) | 12 (9-14) | 7 (5-10) | 3 (2-5) | 3 (2-4) |
| Care home Setting | 2020 | 301 | 318 | 169 | 98 | 30 | 37 | 55 |
| | 5-year average (95% CI) | 324 (283-364) | 402 (355-448) | 273 (216-329) | 429 (360-497) | 205 (167-243) | 156 (121-190) | 167 (153-182) |
| Healthcare Setting | 2020 | 54 | 45 | 34 | 3 | 1 | 4 | 7 |
| | 5-year average (95% CI) | 62 (51-73) | 89 (75-103) | 55 (29-80) | 85 (54-116) | 32 (24-40) | 28 (18-38) | 25 (21-29) |
| Educational Setting | 2020 | 58 | 102 | 53 | 0 | 2 | 3 | 10 |
| | 5-year average (95% CI) | 66 (45-87) | 107 (85-130) | 86 (70-102) | 77 (57-98) | 62 (51-73) | 61 (37-84) | 51 (39-64) |
| Food outlet Setting | 2020 | 29 | 14 | 3 | 1 | 1 | 1 | 6 |
| | 5-year average (95% CI) | 10 (8-12) | 18 (11-25) | 13 (9-17) | 23 (16-30) | 14 (10-17) | 8 (6-11) | 13 (9-17) |
| Custodial Setting | 2020 | 6 | 28 | 1 | 0 | 0 | 1 | 1 |
| | 5-year average (95% CI) | 2 (1-3) | 2 (0-3) | 2 (1-3) | 3 (1-4) | 2 (0-3) | 1 (1-2) | 3 (2-5) |
| Other Settings | 2020 | 17 | 28 | 13 | 7 | 12 | 4 | 12 |
| | 5-year average (95% CI) | 25 (19-32) | 34 (27-41) | 25 (20-31) | 51 (44-57) | 36 (31-41) | 38 (34-42) | 45 (40-50) |
| Laboratory confirmed GI organisms | 2020 | 6023 | 7374 | 3520 | 2859 | 3284 | 4617 | 6205 |
| | 2019 | 5041 | 6499 | 4180 | 8561 | 6967 | 8117 | 9877 |
| | 5 year average (95% CI) | 5016 (4721-5311) | 6471 (5969-6973) | 3943 (3591-4295) | 8345 (7602-9088) | 6553 (5829-7277) | 7879 (7539-8219) | 9303 (8746-9860) |
| | % Change | 20% | 14% | -11% | -66% | -50% | -41% | -33% |

≠ Organisms: Campylobacter, STEC O157, STEC Non-O157, Listeria, non-typhoidal Salmonella, typhoidal Salmonella, Shigella, Norovirus, Cryptosporidium and Giardia

**Supplementary Figure 1 – Laboratory confirmed GI pathogens reported to Public Health England between Week 1 and Week 31 2020 and the 5-year weekly average, by age group and sex**

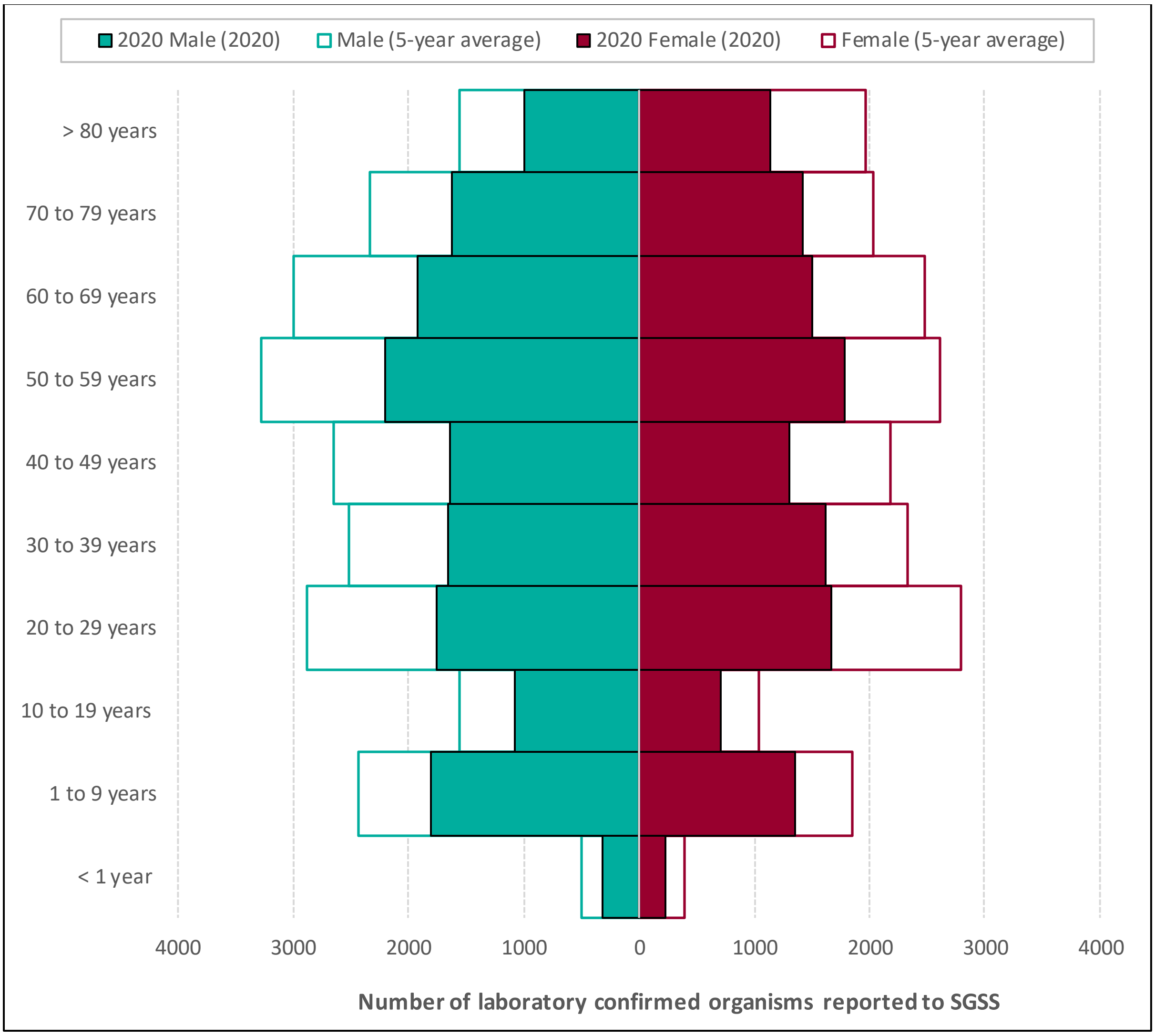

**Supplementary Figure 2 – Laboratory confirmed GI pathogens reported between Week 1 and Week 31 2020 and 5-year weekly average, by COVID-19 outbreak Phase and PHE region**

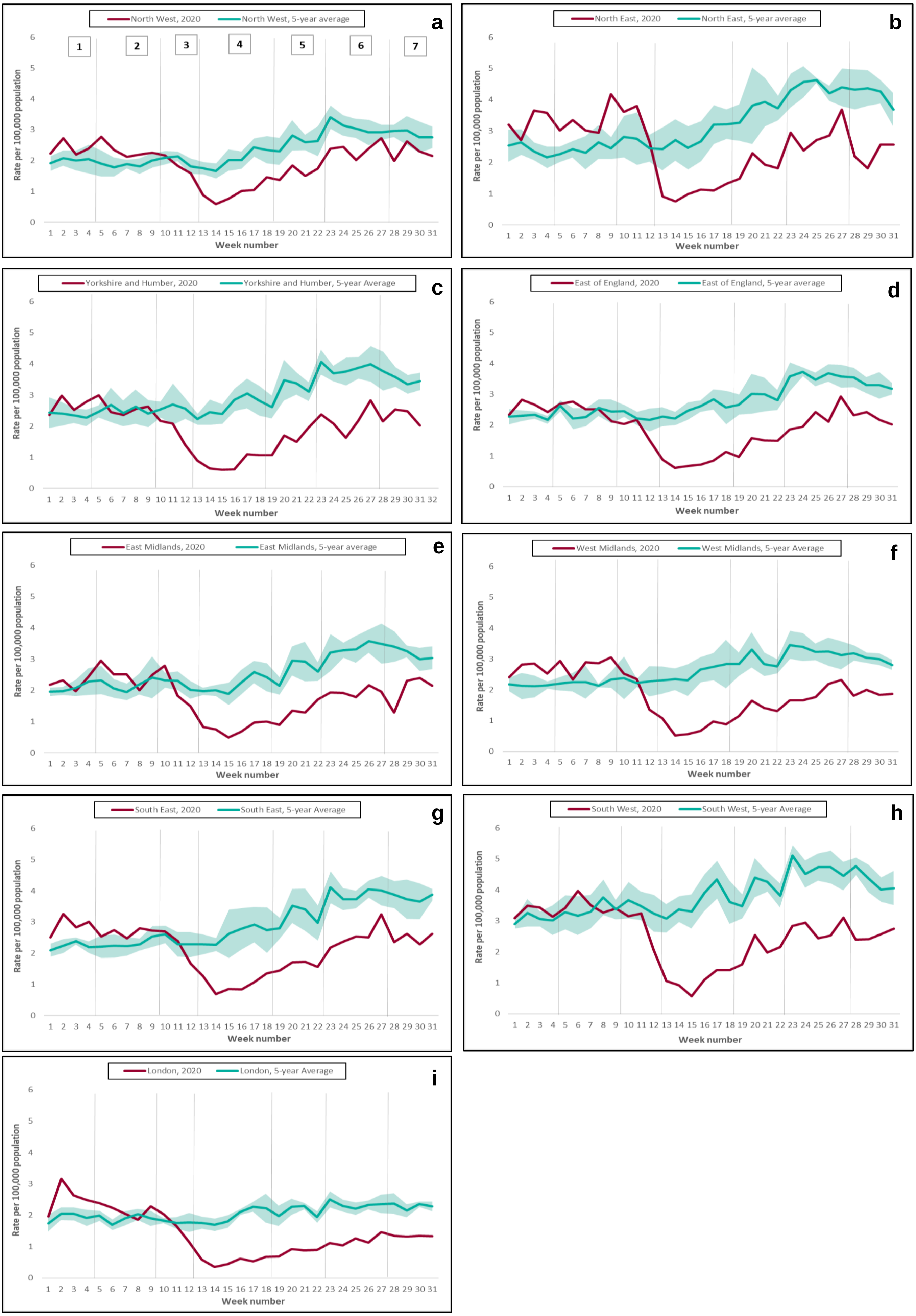

**Supplementary Figure 3 – Correlation between cumulative laboratory confirmed GI diagnoses (percentage relative effect between 2020 and the 5-year average) and cumulative COVID-19 rate for English Upper Tier Local Authorities*, Phase 2 to Phase 7 of the COVID response**

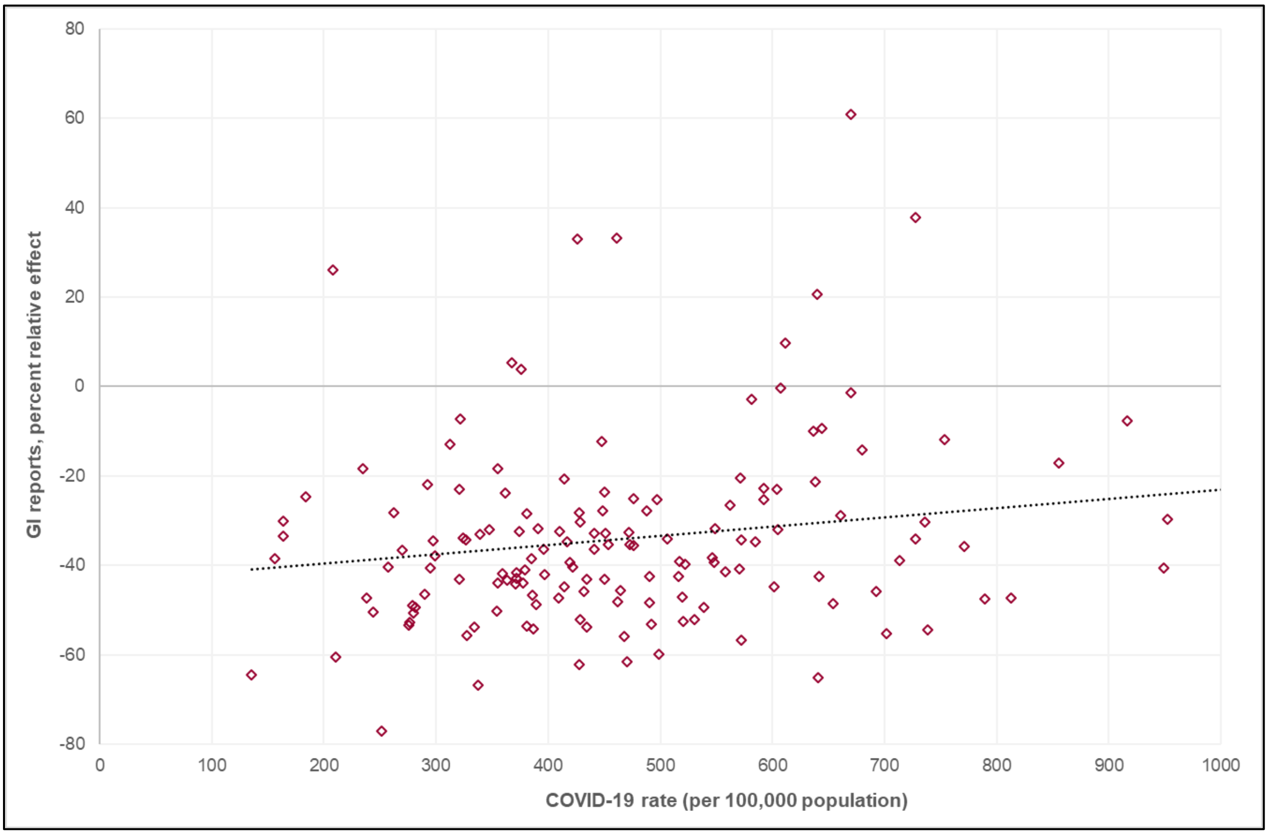


*Graph excludes the Leicester Upper Tier Local Authority due to higher cumulative COVID-19 rate than other local authority areas up to 02 August 2020. Leicester was included in the Pearson’s correlation analysis.

**Supplementary Figure 4 – Daily number of total attendances with a diagnostic code recorded across the Emergency Department Syndromic Surveillance network indicating the seven-day moving average, Phase 1 – 7 of the COVID-19 outbreak response, 2019 and 2020.**

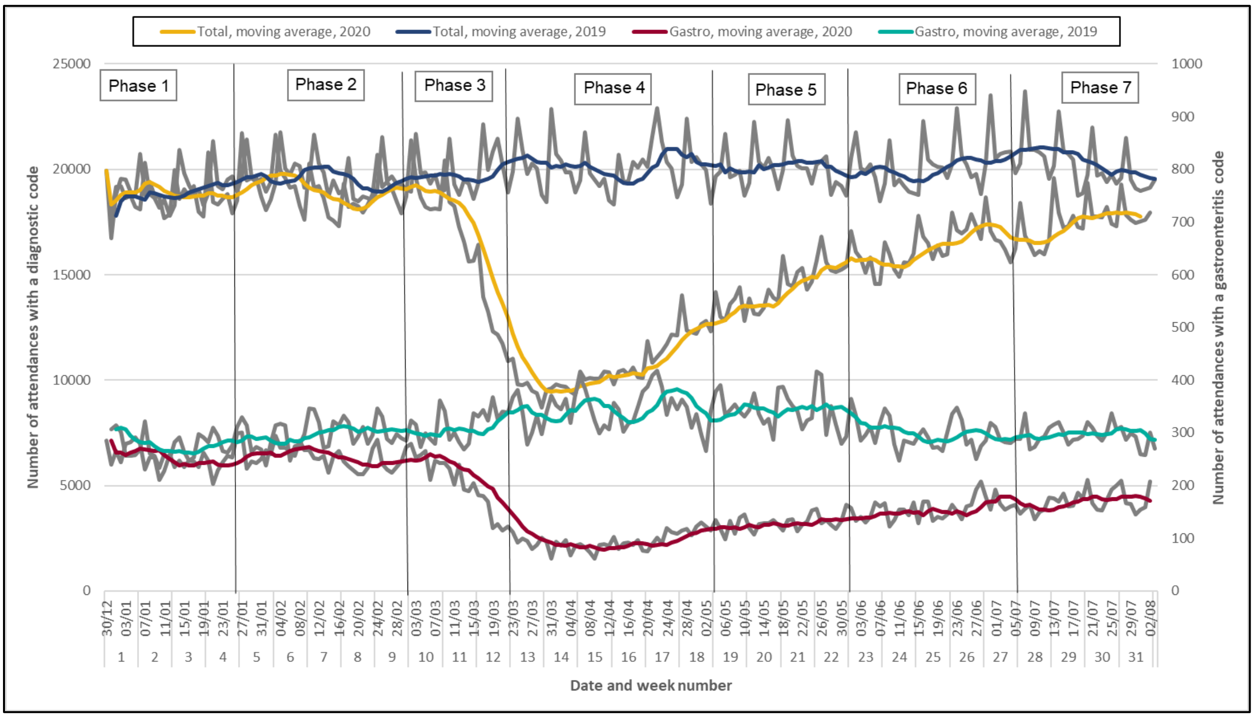

**Supplementary Figure 5 -GP in hours average daily rate by week per 100,000 population for gastroenteritis consultations, COVID-19 outbreak Phase and age group, (a) 2019 and (b) 2020**

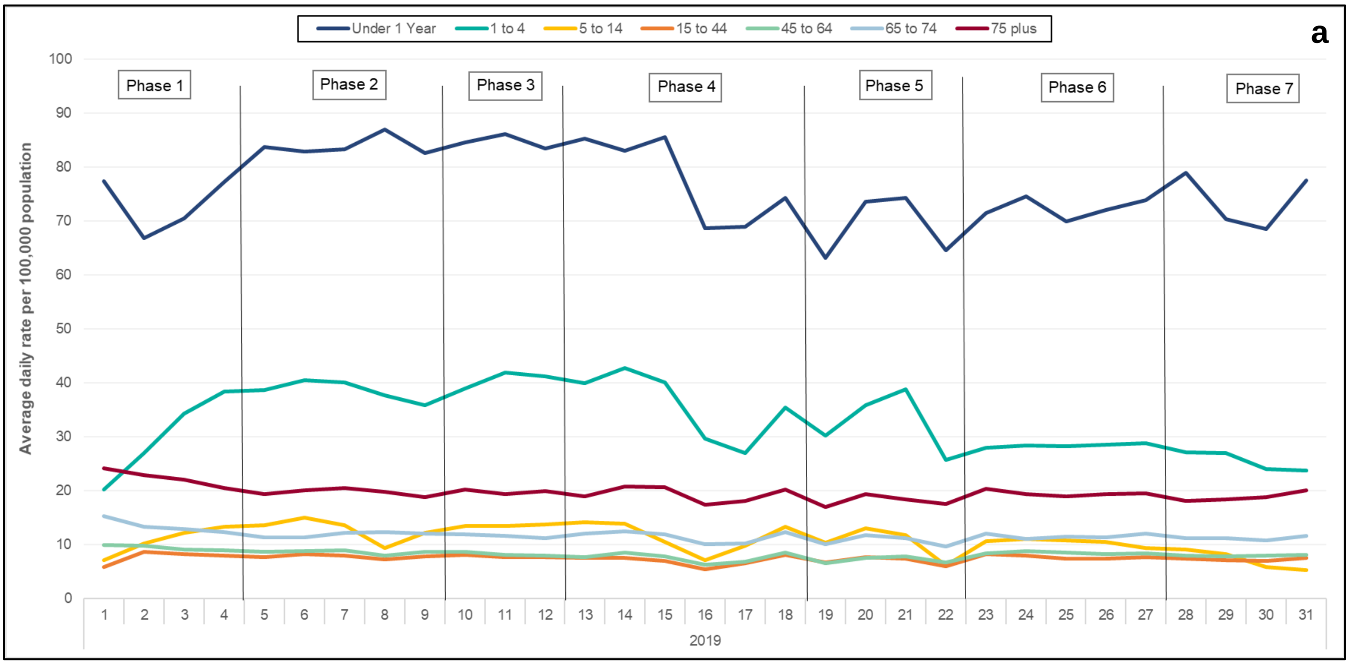


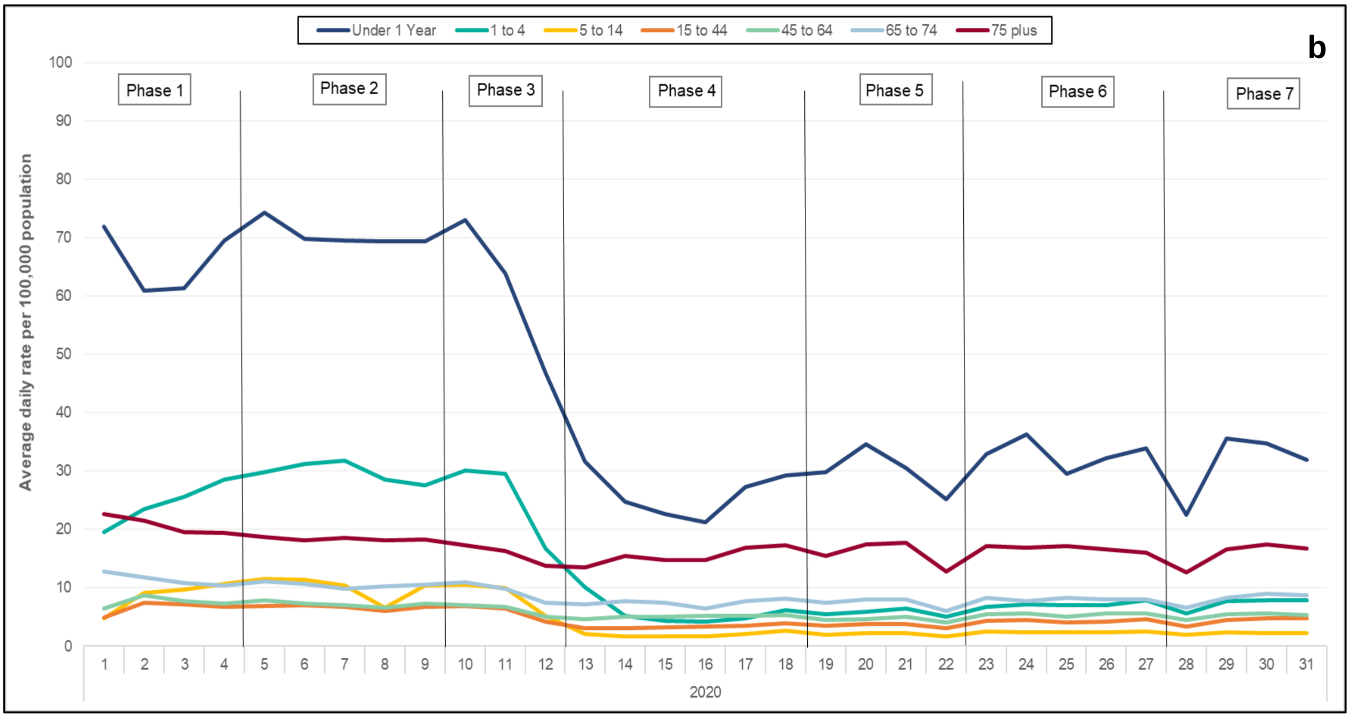

**Supplementary Figure 6 - GP in hours average daily rate by week per 100,000 population for gastroenteritis consultations, COVID-19 outbreak Phase and PHE region, 2019 and 2020**

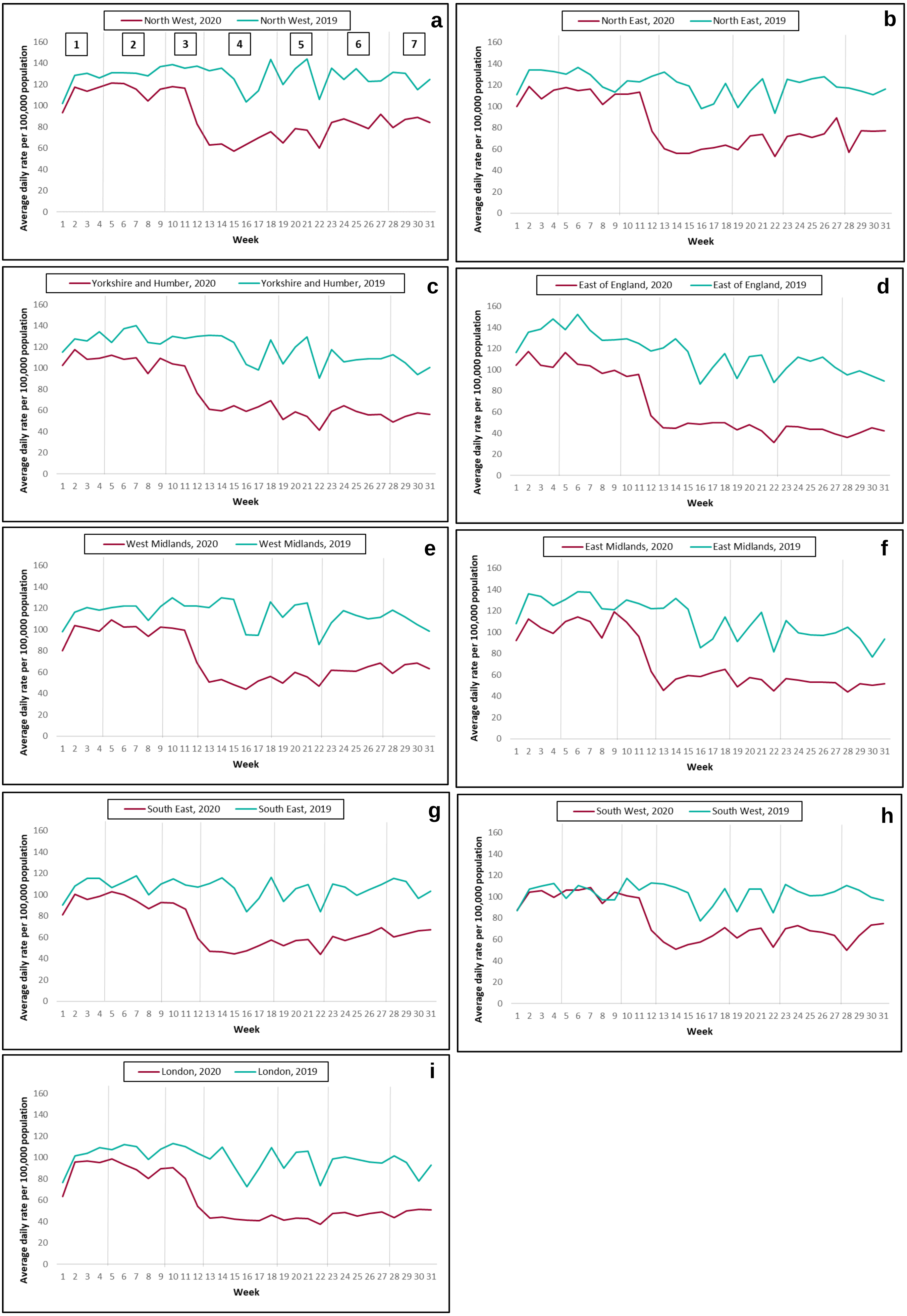

**Supplementary Figure 7 – Daily GP-in hours incidence rate and 7-day moving average (adjusted for weekends and bank holidays) per 100,00 population for (a) vomiting and (b) diarrhoea and daily calls as a percentage of total calls and moving 7-day average for (c) 'Diarrhoea' calls and (d) 'Vomiting' calls**

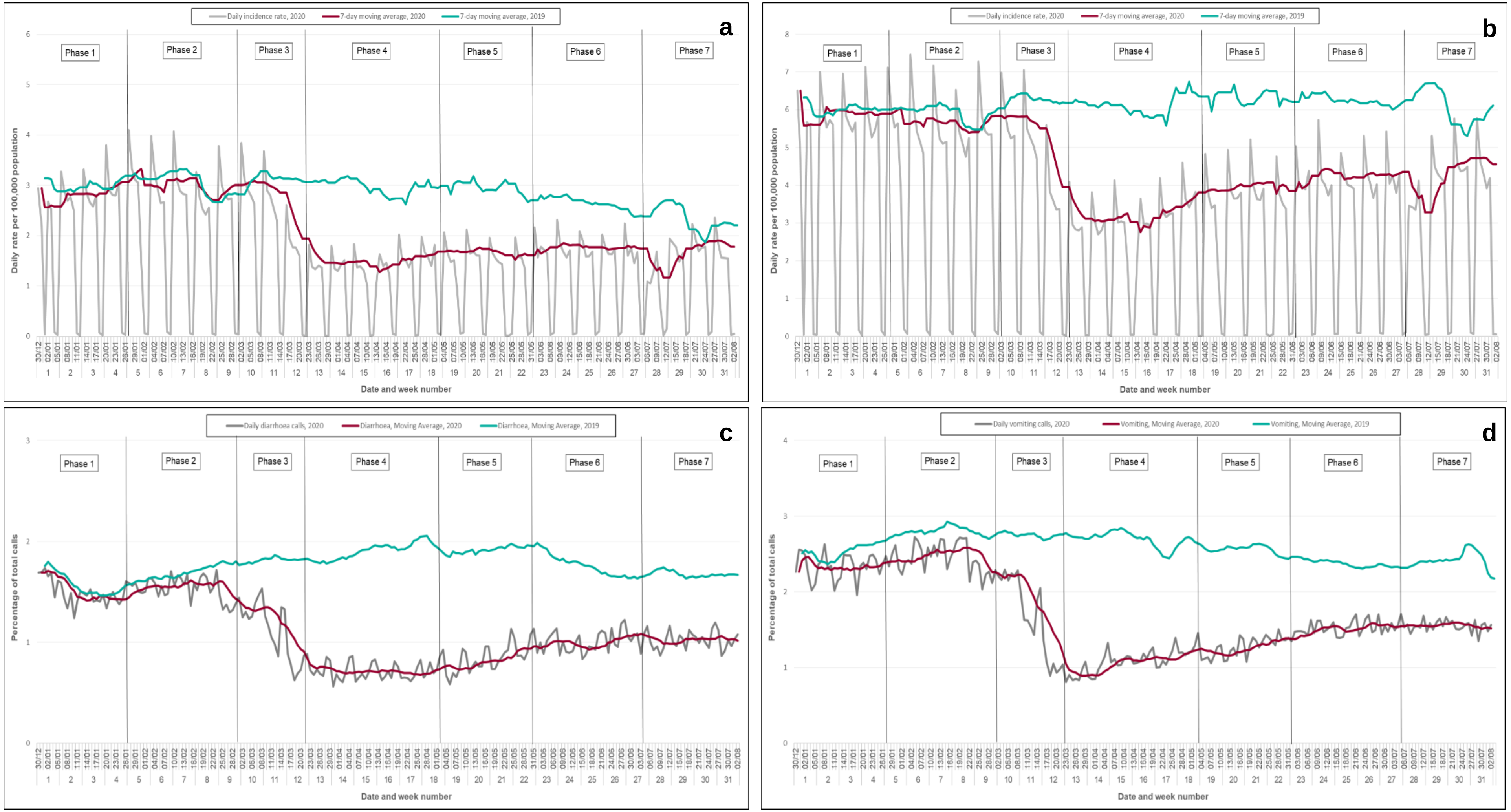

**Supplementary Figure 8 – Google trend searches between 01/01/2017 and 02/08/2020 for (a) protective factors associated with GI illness and (b) key search phrases associated with GI illness, indicating the seven Phases of the COVID-19 response**

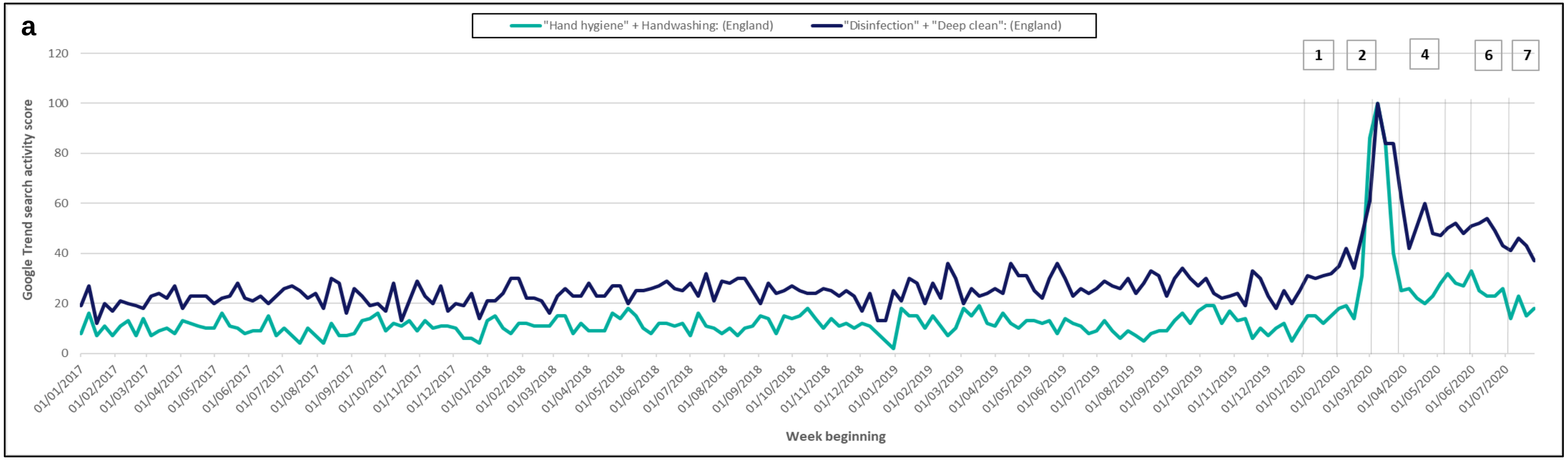


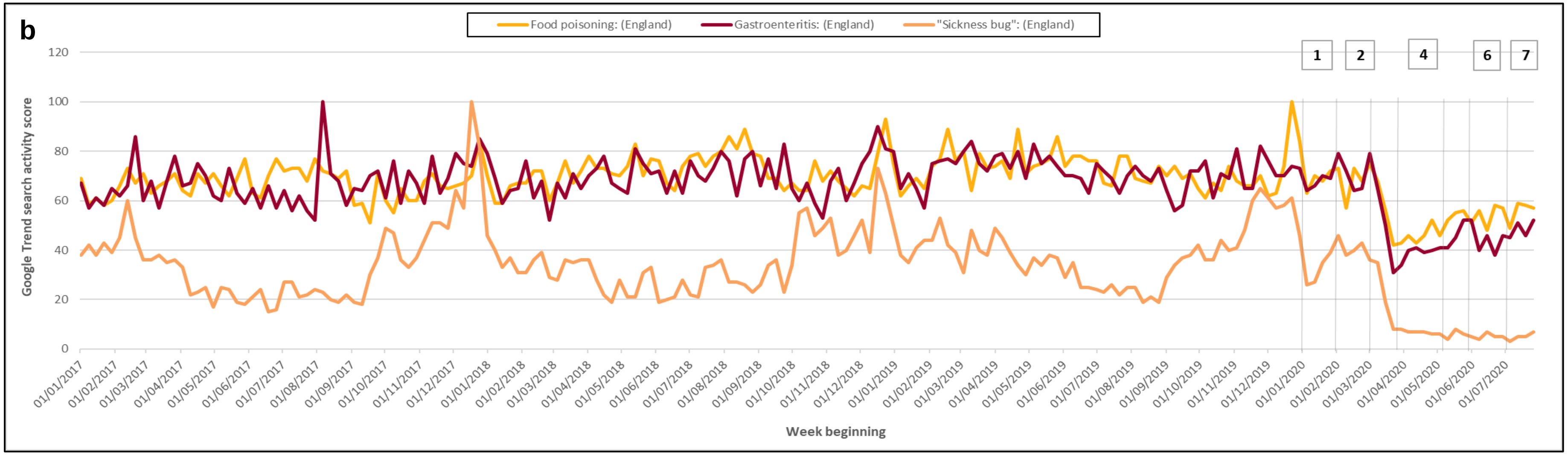
